# Diabetes impact on nigrostriatal vulnerability in Parkinson’s Disease

**DOI:** 10.1101/2025.02.19.25322518

**Authors:** Alice Galli, Cinzia Zatti, Alessandro Lupini, Silvia Paola Caminiti, Andrea Rizzardi, Silvia Lucchini, Francesco Bertagna, Barbara Paghera, Tiago Fleming Outeiro, Daniela Perani, Alessandro Padovani, Andrea Pilotto

## Abstract

Mechanisms underlying the role of diabetes mellitus (DM) as modulator of severity in Parkinson’s Disease (PD) remain unclear. Aim of this study was to investigate the impact of DM on nigrostriatal dopaminergic vulnerability considering two independent cohorts of drug-naïve PD patients (PPMI and DNA studies). Patients with and without DM were first compared and then matched for age, sex, symptoms severity. Differences in ^123^I-FP-CIT binding and molecular connectivity alterations were tested in PD with and without DM within the nigrostriatal pathway. 269 drug-naïve patients were enrolled (PPMI, n=174; DNA, n=95). In both cohorts, patients with DM were older, predominantly male, and exhibited worse non-motor and cognitive symptoms. After severity-matching, patients with DM were found to exhibit more preserved dopamine binding in striatal regions compared to those without DM, together with a lower neural reserve. Patients with DM showed significant dopaminergic connectivity alterations (20%), primarily due to a loss of connectivity (98%). In contrast, patients without DM had more widespread connectivity changes (33%), characterized by both loss (84%) and gained connections (16%). These findings suggest that diabetes directly affects the nigrostriatal network, resulting in less deficits despite similar disease severity, perhaps decreasing the effect of reserve on dopaminergic neurons loss.

## Introduction

Parkinson’s Disease (PD) is a complex and heterogeneous condition in terms of clinical presentation and patterns of progression. Several pre-morbid factors have been advanced to explain this heterogeneity, namely life-style factors ^1^, genetics ^2^, and cardio-vascular factors ^3,4^. Recently, Diabetes Mellitus (DM) has been established as both a risk factor and as an important modulator of alpha-synucleinopathies. The association between DM, worse motor symptoms, and poorer responses to treatments in PD patients was first described in an observational study ^5^ and confirmed by recent studies even in de-novo PD patients ^6,7^. Additionally, DM has been identified as a powerful predictor of cognitive symptoms onset and dementia in PD ^6,8,9^. Common mechanisms underlying this association have been extensively studied. Firstly, DM and PD patients share disruptions in common neural pathways. Indeed, experimental models have demonstrated that prolonged hyperglycemia can be a risk factor for PD pathogenesis and progression, potentially leading to nigrostriatal synaptic dysfunction, increased alpha-synuclein aggregation, and consequent neuroinflammation ^10^ ^11^. Additionally, recent studies have identified possible shared genetic susceptibility mechanisms between DM and PD, putting individuals at risk for both diseases. For instance, the *park7* gene related to PD encodes the DJ-1 protein, which is reduced in pancreatic cells of patients with DM ^12^ and may have deglycase activity^13^. Also, mitochondrial dysfunction and oxidative stress can contribute to onset and progression of both conditions ^14^. Taken together, these findings indicate that DM can increase susceptibility to changes in neural pathways implicated in PD, thus reducing brain’s resilience against neurodegeneration. The concept of brain resilience, originally advanced for brain changes related to normal age or Alzheimer’s disease, has been recently expanded to PD encompassing the concepts of cognitive ^15^and motor reserve ^16,17^. Motor reserve represents an individual’s capacity to maintain motor functionality despite a significant degree of neuropathology in the underlying motor circuitry. The neural basis of motor reserve is the restructuring of both functional and structural brain networks driven by their use throughout life^18^.

Here, we hypothesized that DM may directly affect nigrostriatal pathways, potentially acting as important modulator of brain susceptibility to degeneration of dopaminergic neurons.

Thus, we aimed to conduct an in vivo study using 123I-FP-CIT SPECT imaging in two independent cohorts using a tailored severity-matching procedure in order to establish the role of DM on nigrostriatal system independently from individual motor severity. To achieve this aim, we used complementary analytical strategies, namely the evaluation of regional differences in 123I-FP-CIT binding in nigrostriatal pathway and the assessment of their molecular connectivity alterations in two clinically matched groups of PD patients with and without concomitant DM.

## Materials and methods

### Participants

The study evaluated the relationship between DM and dopaminergic function in two independent cohorts of drug naïve patients: i) Parkinson’s Progression Markers Initiative (PPMI) database, a multicenter, prospective, longitudinal study (www.ppmi-info.org/data); ii) Internal validation data derived from the Single-center Digital Neurodegenerative Assessment (DNA) study, an observational single-center study conducted at the Neurology Unit of the University of Brescia, Italy.

### Multicenter PPMI cohort

Drug-naïve patients were selected from the PPMI dataset according to the following criteria: i) no use of PD medications (i.e., Dopamine agonist, Levodopa); ii) absence of other concomitant neurological disorders; iii) no concomitant anti-depressant medications; iv) availability of clinical assessment at baseline; v) availability of medical history reporting vascular risk factors. Further up-to-date details on general inclusion criteria, protocols and manuals are available online at www.ppmi-info.org/study-design. The institutional review board approved the study at each site, and the participants provided written informed consent.

An age and sex-matched control group (CG) of healthy subjects was selected from PPMI dataset as normative 123I-FP-CIT-SPECT dataset for brain dopaminergic analysis (n=56, age=60.48±10.08, sex (M)=62%).

### Clinical Assessment

Each selected participant underwent a comprehensive evaluation, including a physical examination and assessment of motor, non-motor, and cognitive symptoms. To evaluate the severity of motor symptoms, the MDS Unified Parkinson’s Disease Rating Scale-Part III (MDS-UPDRS-III) was considered ^19^. Non-motor symptoms were evaluated using the MDS-UPDRS-part I ^19^, and the Rapid Eye Movement (REM) Sleep Behavior Disorder Questionnaire (RBDSQ) to assess sleep behavior ^20^. Cardiovascular risk factors were collected from medical history or home therapy, namely type II Diabetes Mellitus (DM), hypertension, hypercholesterolemia, smoking status, and Body Mass Index (BMI)^21^. Global cognition was assessed using the Montreal Cognitive Assessment (MoCA) ^22^. A follow-up clinical assessment was available for a subset of patients in both cohorts.

### 123I-FP-CIT SPECT acquisition and pre-processing

All individuals underwent 123I-FP-CIT SPECT scan at baseline to assess dopaminergic alterations. Brain SPECT imaging was acquired using Siemens or General Electric SPECT tomographs, approximately 3-4 hours after the administration of 123I-FP-CIT tracer. The imaging protocol used for PPMI scans is described in the “SPECT technical operations manual”. Pre-processing of SPECT brain images was conducted using Statistical Parametric Mapping (SPM12), running on Matlab R2022b (MathWorks Inc., Sherborn, MA, USA). First, the origin coordinates of the DAT-SPECT scans were manually set to the anterior commissure. Then, each image was spatially normalized to a high resolution 18 F-DOPA template (http://www.nitrc.org/projects/spmtemplates) ^23^ using the old-normalize function. Parametric binding potential were generated for each subject and voxel-wise using the Image Calculator (ImCalc) function, considering the superior lateral occipital cortex as the reference background region. Only for subsequent voxel-wise analyses, pre-processed parametric images were smoothed with an 8mm FWHM Gaussian kernel to minimize the partial volume effect.

### Single-center DNA cohort

Drug-naïve PD patients were enrolled based on the following exclusion criteria: i) presence of symptoms or features suggesting atypical parkinsonism^24^ ii) dementia; iii) other neurological disorders or medical conditions potentially associated with gait alterations; iv) abnormal MRI; v) bipolar disorder, schizophrenia, history of drug or alcohol abuse or impulse control disorder. The research protocol was approved by the Ethics Committee of the Brescia Hospital, Brescia, Italy (DNA study, NP 1471). Written informed consent was collected from each participant and the study was conducted in accordance with the declaration of Helsinki.

A CG of subjects with a confirmed clinical diagnosis of isolated action or rest tremor syndromes over a 4-year follow-up period and normal 123I-FP-CIT imaging was selected from a normative dataset (see ^25,26^) for brain dopaminergic analysis (n=30, age=72.47±4.51, sex (M)=50%).

### Clinical Assessment

Participants underwent comprehensive clinical evaluations - including a physical examination and assessments of motor, non-motor and cognitive symptoms. The same clinical scales as the PPMI cohort were selected for consistency.

### 123I-FP-CIT SPECT acquisition and pre-processing

Brain SPECT acquisition was performed 3 hours after tracer administration using the Discovery 630, General Electric, Milwaukee, WI. Data were reconstructed by filtered back-projection, with Butterworth 3-dimensional (3D) post-filter (order 10.0; cut-off 0.50 cycle/cm) and corrected for attenuation (Chang’s method coefficient 0.15 cm-1). The same pre-processing pipeline was applied to 123I-FP-CIT SPECT scans as for the PPMI cohort.

### Statistical analysis

Demographic, clinical, and *in vivo* imaging dopaminergic features were compared between PD patients with and without concomitant type II DM (PD-DM and PD-n, respectively), in the PPMI and DNA cohorts. The normality assumption was assessed using the Shapiro-Wilk test and all considered variables met the assumption (p>0.05). Differences in motor, non-motor, and global cognitive symptom severity in PD with and without diabetes was assessed using univariate models, considering age and sex as covariates of nuisance. PD patients with and without DM were then matched 1:1 for sample size, age, sex, and clinical symptoms (i.e., MDS-UPDRS-I, MDS-UPDRS-III, MoCA) separately for the two cohorts, in order to evaluate the impact of DM on dopaminergic alterations independently from parkinsonian symptom severity. The matching procedure utilized the MatchIt library in R version 4.3.1 through the nearest neighbour approach. The same matching procedure was also applied to select a control group for each cohort for subsequent brain dopaminergic analyses.

Significance threshold was set at p<0.05 for all statistical tests. All statistical analyses were performed using JASP (version 0.18.1) and R (version 4.3.1).

### Brain Dopaminergic Analysis

#### Univariate Analysis

Parametric Specific Binding Ratios (SBRs) were extracted following a previously validated pipeline ^25,27,28^ from basal ganglia (i.e., bilateral putamen and caudate) using REX toolbox (https://www.nitrc.org/projects/rex/). ROIs were selected from the Automated Anatomical Labelling (AAL) atlas.^29^ The posterior aspect of the fornix was considered the boundary between the anterior and posterior putamen. SBR in the posterior putamen was considered to calculate asymmetry index (AI) adopting the formula proposed by Walker et al.^30^ :

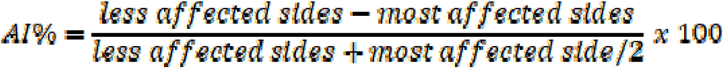

An ANOVA test was conducted to compare the differences in SBR and AI% between PD-DM and PD-n, considering the CG as reference. Additionally, patients with or without concomitant DM were compared using a voxel-wise t-test model in SPM12. The voxel-wise significance threshold was set at p<0.01 (uncorrected) with a minimum cluster extent of 100 voxels.

#### Multivariate Analysis

Nigrostriatal dopaminergic pathway was constructed by selecting specific cortical and subcortical ROIs (i.e., nodes) from the AAL atlas. Only ROIs with mean SBR significantly different from zero in the CG using a one-sample t-test were considered in the following analysis.

### Neural Reserve Calculation

Neural reserve measures were calculated assuming that motor symptoms and dopaminergic denervation are linearly related. The differences between motor symptoms and dopaminergic uptake in striatal regions were used as operational measures of neural reserve using linear regression models, separately for the two included cohorts. Dopaminergic uptake was quantified within i) the whole striatum, ii) within regions resulting from the prior univariate comparison and were considered as the dependent variables. Motor symptoms severity (i.e., MDS-UPDRS-III) was included as the independent variable of interest, controlling for age and AI%. Standardized residuals from this regression were multiplied by −1 to derive W scores^34^. The distribution of W scores was compared between PD-DM and PD-n by means of Welch’s test. Higher W scores reflect greater neural reserve, meaning that a relatively high level of brain damage is tolerated at a given level of motor symptoms.

### Molecular connectivity analyses

Assessment of molecular connectivity between nodes was carried out via Pearson’s correlation analysis. First, SBR values of each included individual were standardized, separately for the two considered cohorts (i.e., PPMI and DNA cohorts), to perform the multivariate analysis in the whole sample. A correlation matrix was computed for each clinical group employing MATLAB’s correlation function to estimate the strength of molecular connectivity between nodes. Nodes formed the nigrostriatal dopaminergic network, and the estimated correlation coefficients were considered as edges ^31^. Fisher’s transformation was applied to each coefficient resulting from the correlation analysis to perform a z-test to assess the significant changes between i) each group of patients and a CG matched for age, sex and sample-size; ii) patients with and without concomitant DM. The significant threshold was set at p<0.05 ^32^. Also, FDR correction for multiple comparisons was applied to obtain more stringent results. The obtained z-scores can be negative or positive, indicating hypo-connectivity or hyper-connectivity, respectively.

To assess and describe molecular network topology in PD-DM and PD-n patients, specific local (nodal) metrics were analyzed using BRAPH software (version 2.0.0.b1) ^33^. Non-parametric tests with n=1000 permutations were carried out to assess differences between groups, which were considered significant for a two-tailed test of the null hypothesis at p<0.05. In detail, we selected the binary undirected density (BUD) method to assess:

- *Degree:* the average of the shortest inverse distances between a node and all other nodes in the network, meaning that the nodal degree is the total number of the node’s connections;
- *Path length*: the average distance from a given node to all other nodes, which is inversely proportional to the connection strength;
- *Local efficiency*: the average of the shortest inverse distances among nodes within the local subgraph (all neighboring nodes of a given node and all edges among them); this measure is useful to assess the communication efficiency between each node and its immediate neighbors.

## Data availability

The data that support the findings of this study are available from the corresponding author, upon reasonable request.

## Results

### Baseline Clinical Assessment

In the multicenter PPMI cohort, out of 497 PD participants enrolled, 186 drug-naïve PD individuals met the selection criteria for the presence of vascular risk factors and glucose levels at baseline. Eleven out of the 186 patients showed artifacts in dopaminergic imaging and were subsequently excluded from further analyses. The total sample included 174 patients, namely 56 with and 118 PD patients without DM (see the flowchart-**Supplementary Figure 1**). At baseline, PD-DM were found to be older (p=0.009), have a higher percentage of males (83%, p=0.002), and exhibit worse non-motor (MDS-UPDRS-I, p=0.011) and cognitive symptoms (MoCA, p=0.035) than those with PD-n (**Supplementary Table 2**).

The single-center DNA cohort included 110 drug-naïve PD patients. Among them, five patients were excluded for the absence of clinical data, thus obtaining an eligible sample of 105 patients including 30 patients with PD-DM and 80 patients with PD-n. Ten PD patients were further excluded due to artifacts in dopaminergic imaging, resulting in a final sample of 95 patients, namely 30 and 65 PD patients with and without DM, respectively (**Supplementary Figure 1**). At baseline, patients with DM were slightly older (p=0.060), had a higher percentage of males (73%, p=0.035), and exhibited worse cognitive symptoms (MoCA, p=0.039) compared to patients without DM (**Supplementary Table 2**).

#### Matching Procedure

For subsequent analyses, a 1:1 matching of PD-n with PD-DM patients based on age, sex, motor, and non-motor symptoms was performed in both cohorts. As expected, after the matching procedure, PD-DM and PD-n were comparable in both the cohorts under consideration. Patients’ demographics and clinical features after the matching procedure are shown in **Table 1**.

**Table 1.** Demographic and clinical characteristics of the included patients after the matching procedure.

|  | PD-DM | PD-n | p-value |
| --- | --- | --- | --- |
| **PPMI cohort (n)** | 56 | 56 | - |
| Age (mean ± SD) | 64.73±8.61 | 62.99±10.97 | 0.513 |
| Sex (F/M) | 9/47 | 7/49 | 0.589 |
| MDS-UPDRS-I (mean ± SD) | 5.88±4.29 | 5.05±2.69 | 0.460 |
| MDS-UPDRS-III (mean ± SD) | 22.61±2.14 | 21.07±8.94 | 0.512 |
| MoCA (mean ± SD) | 26.21±2.91 | 27.05±2.37 | 0.150 |
| **DNA cohort (n)** | 30 | 30 | - |
| Age (mean ± SD) | 69.88±7.27 | 69.27±5.76 | 0.722 |
| Sex (F/M) | 8/22 | 11/19 | 0.405 |
| MDS-UPDRS-I (mean ± SD) | 6.17±4.90 | 5.31±4.98 | 0.508 |
| MDS-UPDRS-III (mean ± SD) | 12.93±9.21 | 14.17±7.07 | 0.563 |
| MoCA (mean ± SD) | 25.70±2.84 | 26.07±1.64 | 0.543 |

*Abbreviations: PD, Parkinson's Disease; DM, Diabetes Mellitus; MDS-UPDRS, Movement Disorder Society Unified Parkinson's Disease Rating Scale; MoCA = Montreal Cognitive Assessment.*

### Brain Dopaminergic Activity

#### Univariate Analysis

The comparison between matched PD-n and PD-DM groups in dopaminergic binding is shown in **Figure 1** and **Table 2**.

**Figure 1.**
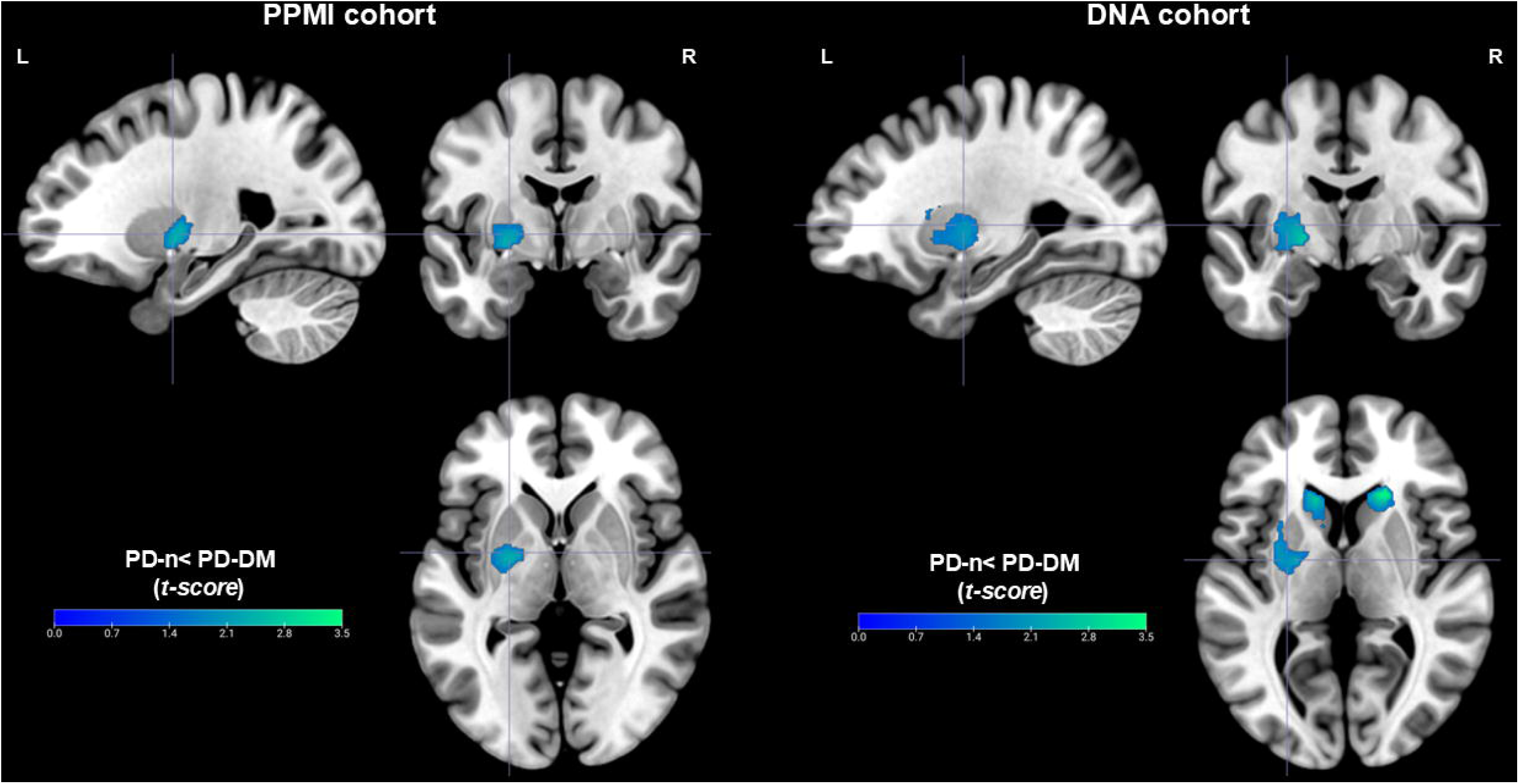
Voxel-wise SPM comparison between PD-n and PD-DM. Significant decreased dopaminergic binding in PD-n as compared to PD-DM in basal ganglia overlaid on a T1-standard template. Results are represented separately for DNA and PPMI cohorts. Abbreviations: PD, Parkinson’s Disease; DM, Diabetes Mellitus; L, Left; R, Right.

**Figure 2.**
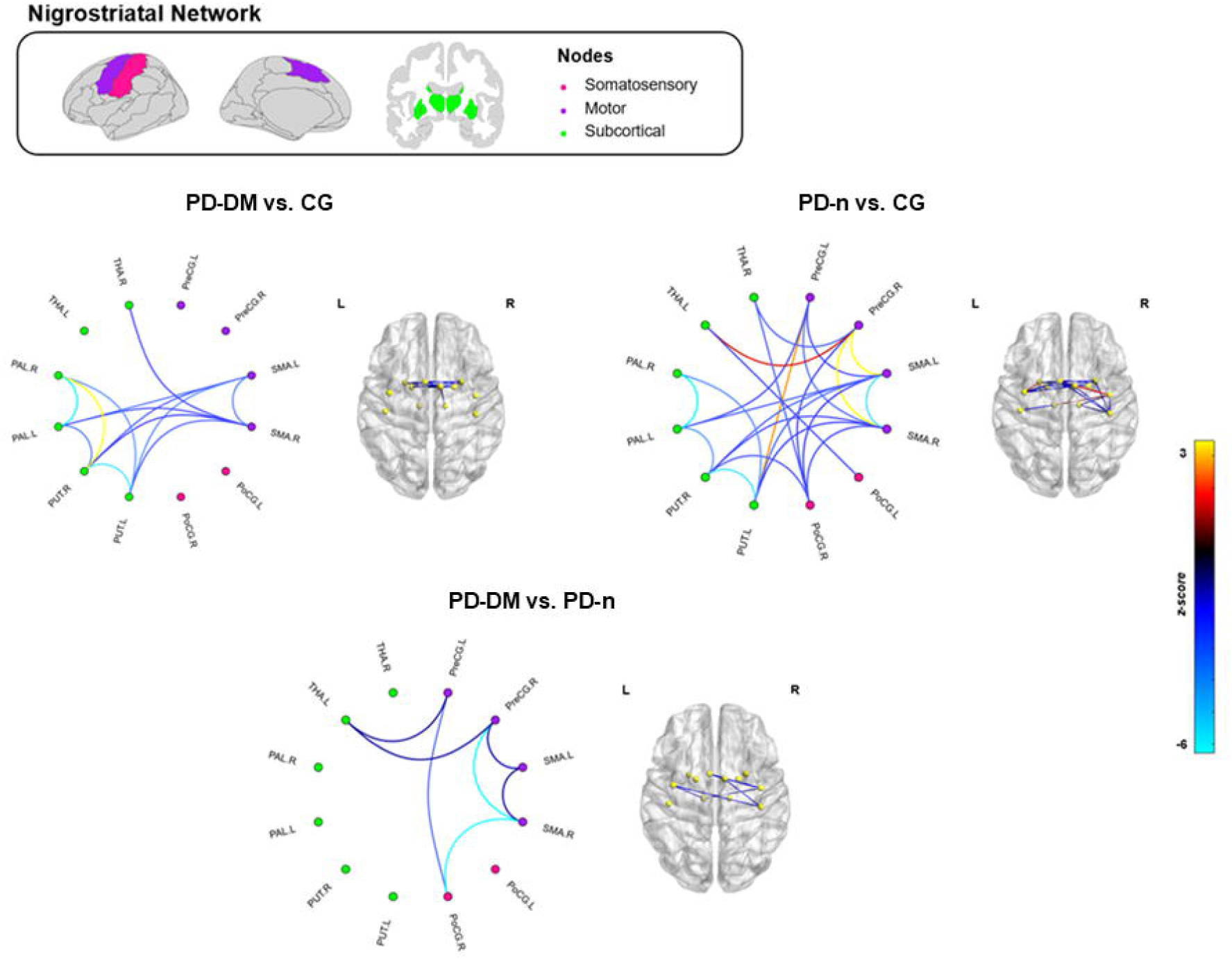
Nigrostriatal Dopaminergic Network Analysis. Network plots represent the significant differences obtained comparing correlation (p<0.05). Connectivity losses are shown in cyan-blue, while gained connectivity is shown in yellow-red. Unchanged connections are hidden. 3D brain renders showed connectivity changes, with yellow circles representing networks’ nodes. Abbreviations PD, Parkinson’s Disease; DM, Diabetes Mellitus; L, Left; R, Right; THA, Thalamus; PAL, Globus Pallidus; PUT, Putamen; SMA, Supplementary Motor Area; PoCG, Postcentral Gyrus; PreCG, Precentral Gyrus.

**Table 2.** Dopaminergic binding in basal ganglia at baseline.

|  | CG | PD-DM | PD-n | p-value |
| --- | --- | --- | --- | --- |
| **PPMI cohort (n)** | 56 | 56 | 56 | - |
| Putamen L | 2.25±0.39 | 1.44±0.52 | 1.28±0.35 | 0.024<sup>a,b,c</sup> |
| Putamen R | 2.06±0.35 | 1.34±0.46 | 1.28±0.34 | <0.001<sup>a,b</sup> |
| Caudate L | 1.45±0.34 | 0.99±0.41 | 0.96±0.29 | <0.001<sup>a,b</sup> |
| Caudate R | 1.44±0.36 | 0.95±0.37 | 0.95±0.28 | <0.001<sup>a,b</sup> |
| Asymmetry Index (%) | 7.97±7.48 | 16.61±10.27 | 12.23±13.33 | <0.001<sup>a,c</sup> |
| **DNA cohort (n)** | 30 | 30 | 30 | - |
| Putamen L | 2.48±0.34 | 2.06±0.38 | 1.88±0.48 | <0.001<sup>a,b,c</sup> |
| Putamen R | 2.25±0.29 | 1.85±0.34 | 1.72±0.46 | <0.001<sup>a,b</sup> |
| Caudate L | 1.58±0.34 | 1.55±0.33 | 1.39±0.38 | 0.059 |
| Caudate R | 1.56±0.29 | 1.49±0.34 | 1.36±0.39 | 0.082 |
| Asymmetry Index (%) | 5.99±5.27 | 23.04±12.99 | 21.44±15.11 | <0.001<sup>a,b</sup> |

*Abbreviations: PD, Parkinson's Disease; DM, Diabetes Mellitus; L, Left; R, Right. <sup>a</sup>= PD-DM <sup>a</sup>= PD-DM < CG; <sup>b</sup>= PD-n < CG; <sup>c</sup>= PD-n < PD-DM. Post-hoc comparisons are Bonferroni corrected for multiple comparisons. Data are expressed as mean SBR ± st.deviation.*

In the multicenter PPMI cohort, both PD-DM and PD-n showed significant dopaminergic reductions within bilateral putamen (p<0.001) and caudate (p<0.001) as compared to CG. The SBR direct comparison showed more preserved mean SBR values in the left putamen (p=0.023) in PD-DM as compared to PD-n with similar cognitive and motor severity. In the voxel-wise direct comparison, PD-DM exhibited more preserved dopamine uptake in left putamen and pallidum (p<0.01 at peak level and p<0.05 at cluster level; cluster listed in **Supplementary Table 3**).

In the single-center DNA cohort, both PD-DM and PD-n exhibited reduced mean SBR in bilateral putamen (p<0.001) than CG- as expected. The SBR direct comparison confirmed higher mean SBR values in left putamen in PD-DM patients than PD-n (p=0.047). The voxel-wise analysis showed higher dopamine uptake in bilateral putamen and caudate in PD-DM vs. PD-n (p<0.01 at peak level and p<0.05 at cluster level, cluster listed in **Supplementary Table 3**).

#### Neural Reserve Measures

Welch’s test revealed significant differences between PD-DM and PD-n for the whole striatum W scores (PPMI, p=0.125; DNA, p=0.031) and the left putamen W scores (PPMI, p=0.034; DNA, p=0.048). In details, PD-DM showed lower W scores than PD-n, suggesting a reduced neural reserve.

#### Multivariate Analysis

Bilateral putamen, globus pallidus, thalamus, postcentral and precentral gyri, and supplementary motor area were considered as nigrostriatal nodes. Middle frontal gyrus showed mean SBR comparable to zero in CG and, thus, was excluded from further analyses. PD-DM showed significantly altered connectivity in the 20% of nigrostriatal nodes, mostly characterized by connectivity losses (i.e., z-scores <0; 92% of altered connections). PD-DM showed short-distance connectivity alterations between putamen and globus pallidus and their contralateral homologous nodes. Also, the right putamen showed gained connections (i.e., z-scores>0) with the right globus pallidus. Long-distance connectivity alterations were found between subcortical nodes and supplementary motor area. Only connectivity losses survived the FDR correction for multiple comparison.

PD-n showed statistically significant altered connections in the 33% of nigrostriatal nodes. Of note, PD-n showed prevalent connectivity losses (i.e., 84% of altered connections) involving short-distance between putamen and globus pallidus, as well as long-distance connections between subcortical, somatosensory, and motor nodes. PD-n exhibited gained connectivity (i.e., 16% of altered connections) involving long-distance connections between putamen and precentral gyrus, as well as between thalamus and precentral gyrus. Short-distance gained connections were found between motor nodes. Following FDR correction, putamen and globus pallidus showed connectivity losses with their contralateral homologous and to supplementary motor area, while gained connectivity characterized left putamen and precentral gyrus.

The direct comparison revealed significantly decreased connectivity between left thalamus and frontal nodes, as well as within frontal nodes (i.e., precentral gyrus and supplementary motor area) in PD-DM as compared to PD-n.

#### Graph Theory Metrics

Network analyses were conducted on binary undirected graphs, with densities fixed at 20%. Considering all nodal metrics, both PD-DM and PD-n showed significant differences as compared to CG. Of note, reduced nodal *degree* (i.e, reduced number of connections starting from the node) was found in subcortical and motor nodes both in PD-DM and PD-n as compared to CG. The *path length* was significantly increased (i.e., lower strength in connections starting from the node) in the globus pallidus both in PD-DM and PD-n than CG. The *local efficiency* was significantly decreased in the globus pallidus in PD-DM vs. CG, while it was increased in globus pallidus and putamen in PD-n vs. CG.

The direct comparison between the two groups of PD patients revealed significantly decreased nodal *degree* (i.e, reduced number of connections starting from the node) in left putamen and globus pallidus in PD-DM as compared to PD-n. The *path length* was significantly increased in the right globus pallidus, and supplementary motor area in PD-DM vs. PD-n. The *local efficiency* was significantly reduced in the left putamen and globus pallidus in PD-DM when compared to PD-n.

## Discussion

Diabetes is a known risk factor and modulator of PD but the mechanisms underlying this association are still under-investigated. These findings demonstrated in two independent cohorts that PD with concomitant DM exhibit a distinct pattern of nigrostriatal dopaminergic binding and connectivity changes compared to PD patients without DM matched for motor and non-motor symptoms severity. These results provide new in-vivo evidence that DM significantly affects nigrostriatal dopaminergic network vulnerability in PD.

In the present study, the baseline clinical assessment revealed that PD patients with DM had worse non-motor and cognitive symptoms compared to PD, in both the tested cohorts. This finding is consistent with previous research suggesting that DM may negatively affect cognitive functions in patients with PD, resulting in an increased risk of developing mild cognitive impairment over time ^35,36^. This faster cognitive deterioration may be due to mechanisms other than disease-specific neurodegeneration, such as insulin resistance ^36^, , but also mitochondrial dysfunction ^35^ or neuroinflammation ^35^, as demonstrated for other neurodegenerative conditions ^37,38^. These factors could be potentially influenced also by the large and significant overlap with other comorbidities including cardiovascular diseases, and metabolic syndrome^3,4^.

Growing literature based on animal and *in vitro* models suggests that DM may induce neurodegeneration by making the nigrostriatal pathway more vulnerable and less resilient ^7,39^. For instance, dysregulation of glucose metabolism can appear years before motor symptoms, suggesting the crucial role of hyperglycemia in facilitating the onset of PD ^40^. Notably, previous in vitro and animal studies have described the presence of insulin receptors in the basal ganglia and substantia nigra ^41^, as well as the key role of insulin in regulation of the dopaminergic transmission^42,43^.

In order to evaluate the specific impact of diabetes on dopaminergic networks independently from direct (and indirect) relationship with motor severity, an accurate matching for age, sex, motor, cognitive and autonomic symptoms was applied in both cohorts. Our results revealed *in vivo* differences in dopaminergic binding and connectivity between matched PD-n and PD-DM. First, PD-DM patients showed a higher dopamine binding in basal ganglia, in comparison to the PD-n, meaning that PD-DM need a lower level of dopaminergic alteration to reveal comparable levels of disease severity (assessed/matched using MDS-UPDRS-I and III). This result was further supported by the voxel-wise results, demonstrating higher dopamine binding in PD-DM in basal ganglia in patients matched for severity in both cohorts. Notably, a previously validated method^34^ was applied to obtain standardized individual differences between predicted and observed dopaminergic deficits within the whole striatum and the left putamen. PD-DM showed lower W scores than PD-n, suggesting that less dopaminergic dysfunction is tolerated at comparable levels of motor deficits in the former group. The consistency between the two cohorts provides robustness to the findings, as they differ for design (multi-center vs single-center) and recruitment strategies, with the PPMI cohort represented by younger but generally more severe patients (21.72±10.1 vs 13.55±8.2 MDS-UPDRS-III for PPMI and DNA cohorts respectively). Of note, only in the PPMI cohort both PD-DM and PD-n showed significant dopaminergic alterations in both the putamen and caudate.

The multivariate connectivity analysis provided further evidence of the DM impact on nigrostriatal network integrity, which is a core feature of PD^24^. Compared to age and sex-matched controls, PD-DM showed a less prominent derangement within nigrostriatal pathway (20% of total connections are altered) as compared to PD-n (33%), supporting the univariate analysis findings. Of note, connectivity alterations were characterized by prevalent loss of connectivity in PD-DM (92% of altered connections) involving both short-distance subcortical and long-distance connections. Furthermore, PD without DM showed increased connectivity in the 16% of altered connections, namely putamen and thalamus projections to frontal nodes. The presence of increased connectivity, involving regions with 123I-FP-CIT bindings known to be reduced in PD, is a consistent finding ^31^described in a growing amount of literature as an early phase response to counterbalance the neurodegenerative processes^25,44^ in both Alzheimer’s Disease and Parkinson’s Disease^45,46^. The advanced hypotheses suggest an ongoing compensatory or maladaptive mechanism due to underlying pathological process^31,47^.

The direct comparison between PD with and without concomitant DM demonstrated the presence of a more decreased connectivity in PD-DM affecting thalamus and frontal nodes, crucial regions for motor and non-motor symptoms onset. The derangement of connections involving the supplementary motor area may contribute to a reduced motor adaptability often reported in patients with PD and concomitant cardiovascular comorbidities^3,48^. The more pronounced loss of long-distance connections in PD-DM may suggest worse compensatory mechanisms within nigrostriatal network. Additional graph theory metrics were investigated to better describe the topography of network reconfiguration driven by diabetes mellitus. As expected, some common alterations were found in the comparison of each group of PD patients with controls. For instance, significantly reduced number of connections starting from basal ganglia towards frontal nodes were found in both PD-DM and PD-n as compared to CG. Consistently, in both PD groups, connections starting from the globus pallidus were weaker than in the control group. Interestingly, the *local efficiency* (i.e., a measure of communication efficiency) was found to be reduced in basal ganglia in PD-DM but increased in PD-n, aligning with the presence of increased connectivity characterizing PD without DM. The direct comparison between PD-DM and PD-n further supports the greater proportion of decreased connectivity driven by DM resulting in decreased and less efficient connections starting from crucial nodes within the nigrostriatal pathway. The complex interplay between metabolic dysregulation and neurological health has been described in a previous study conducted in individuals without PD, showing functional resting-state connectivity abnormalities in fronto-striatal circuits (i.e., reduced connectivity between striatum and motor cortices) in patients with DM.^49^.

Taken together, these findings highlighted the role of DM as an important modifier of brain vulnerability and resilience in PD, especially by impairing the dopaminergic networks to adapt to the progressive loss of nigrostriatal neurons.

These results have several research and clinical implications. First, findings highlight the importance of early screening and management of vascular and metabolic risk factors in the general population, and specifically in subjects at-risk for PD. The increased risk of developing PD in subjects with diabetes prompted its inclusion in the prodromal PD criteria. ^50–52^. Still, insulin resistance and diabetes screening are often neglected in neurological settings potentially limiting an important early window of intervention. Patients with both PD and DM are known to experience faster progression of cognitive symptoms, gait disturbances, and postural instability^35^. Studying mechanisms underlying the association between PD and DM may also raise the possibility of new therapeutic targets acting both on metabolic changes and neurodegeneration. Insulin has demonstrated protective effects on dopaminergic neurons in experimental models^51^ and Glucagon-like peptide 1 (GLP-1) agonists have reported a significant neuroprotective effect in preclinical studies of PD by reducing neuroinflammation and oxidative stress ^53,54^. The present findings indicate a specific negative role of DM- on nigrostriatal system in PD and support potentially GLP-1 treatments–, which should be definitively further explored in early disease phases as target engagement in on-going clinical trials in early PD. ^55,56^

### Limitations and future directions

We acknowledge some limitations of the study. First, the absence of correction for partial volume effects might have led to subtle localization biases. However, we used anatomical atlas for ROI segmentation and used non-smoothed images for analyses^25,28^. Second, we use dopaminergic imaging as proxy of motor reserve in PD but the integration with a clinical scale evaluating other factors impacting motor reserve will be important and has been included in ongoing prospective studies^57^. Third, the work could not evaluate prospective changes in insulin/glucose level and the potential impact of pharmacological drugs, which potentially act against neurodegeneration beyond their impact on metabolic control.^35^ Limitation notwithstanding, this study has several strengths. First, we only enrolled newly diagnosed PD patients who are not on anti-parkinsonian medication. Second, we included two different and independent cohorts to improve the generalizability of the results in both monocentric and multicenter cohorts using different dopaminergic imaging scans. Third, the application of advanced connectivity analyses confirmed and expanded the findings using ROI-Based analyses, suggesting a mechanism of DM-related connectivity alterations beyond the impact on dopaminergic levels.

### Conclusions

Overall, the study demonstrated the modulating effect of DM on the nigrostriatal system in two independent PD cohorts and highlighted the relevance of metabolic changes on compensatory mechanisms and resilience to neurodegeneration within the PD spectrum.

## Supplementary material

Supplementary material is available at *Brain* online.

