## Supplementary Materials for "Diabetes impact on nigrostriatal vulnerability in Parkinson’s Disease"

**Supplementary Figure 1-** Flowchart representing patients’ enrollment and the final sample of the two independent cohorts.

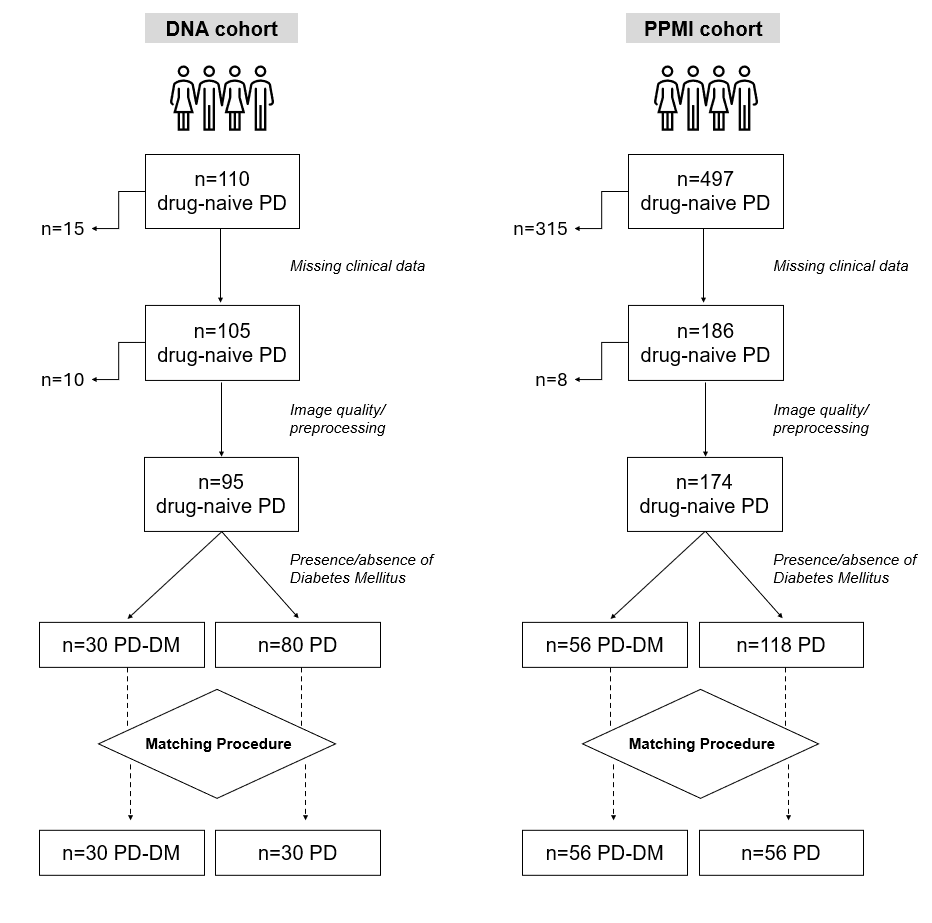
 On the left side the number of patients excluded in each step as reported. *Abbreviations: PD, Parkinson’s Disease; DNA, Digital Neurodegenerative Assessment cohort from the University of Brescia; PPMI, Parkinson’s Progression Markers Initiative cohort; DM, Diabetes Mellitus.*

**Supplementary Table 1 –** Vascular Risk Factors in the two considered cohorts.

|  | PD-DM | PD | Tot. | p-value |
| --- | --- | --- | --- | --- |
| **PPMI cohort (n)** | 56 | 118 | 174 | - |
| Diabetes Mellitus (%) | 56 (100%) | 0 (-) | 56 (32%) | **<0.001** |
| Hypertension (%) | 28 (50%) | 27 (23%) | 55 (32%) | **<0.001** |
| Hypercholesterolemia (%) | 15 (27%) | 8 (7%) | 23 (13%) | **<0.001** |
| Smoking Status (%) | 4 (33%) | 14 (26%) | 18 (10%) | 0.629 |
| Body Mass Index (mean ± SD) | 28.72±5.89 | 26.60±4.47 | 27.29±5.05 | **0.010** |
| **DNA cohort (n)** | 30 | 45 | 75 |  |
| Diabetes Mellitus (%) | 30 (100%) | 0 (-) | 30 (40%) | **<0.001** |
| Hypertension (%) | 20 (66%) | 15 (33%) | 35 (47%) | **0.005** |
| Hypercholesterolemia (%) | 13 (60%) | 18 (29%) | 31 (45%) | **0.007** |
| Smoking Status (%) | 2 (7%) | 8 (18%) | 10 (13%) | 0.121 |
| Body Mass Index (mean ± SD) | 28.07±4.04 | 24.75±3.85 | 26.05±4.23 | **<0.001** |

*Abbreviations: PPMI-PD, Parkinson’s Progression Markers Initiative cohort; DMA-PD, University of Brescia internal cohort.*

**Supplementary Table 2-** Demographic and clinical characteristics of the included patients.

|  | PD-DM | PD-n | p-value |
| --- | --- | --- | --- |
| **PPMI cohort (n)** | 56 | 118 | - |
| Age (mean ± SD) | 64.73±8.61 | 60.31±10.99 | 0.009 |
| Sex (F/M) | 9/47 | 47/71 | 0.002 |
| MDS-UPDRS-I (mean ± SD) | 5.88±4.29 | 4.46±2.91 | 0.011 |
| RBDSQ (mean ± SD) | 4.77±3.71 | 4.92±2.95 | 0.771 |
| MDS-UPDRS-III (mean ± SD) | 22.61±2.14 | 20.54±11.16 | 0.191 |
| MoCA (mean ± SD) | 26.21±2.91 | 27.29±2.14 | 0.035 |
| **DNA cohort (n)** | 30 | 65 | - |
| Age (mean ± SD) | 69.88±7.27 | 66.715±6.85 | 0.060 |
| Sex (F/M) | 8/22 | 22/23 | 0.035 |
| MDS-UPDRS-I (mean ± SD) | 6.17±4.90 | 4.98±4.41 | 0.280 |
| RBDSQ (mean ± SD) | 2.71±3.43 | 3.18±3.29 | 0.585 |
| MDS-UPDRS-III (mean ± SD) | 12.93±9.21 | 13.07±6.59 | 0.942 |
| MoCA (mean ± SD) | 24.96±1.97 | 26.83±2.02 | 0.039 |

*Abbreviations: p-value was adjusted for age and sex via ANCOVA; PD, Parkinson’s Disease; DM, Diabetes Mellitus; MDS-UPDRS, Movement Disorder Society Unified Parkinson’s Disease Rating Scale; MoCA = Montreal Cognitive Assessment.*

**Supplementary Table 3-** Significant voxel-wise differences between PD-DM and PD-n within the two independent cohorts.

*Anatomical labeling was obtained using AAL atlas version 2.*

|  |  | **MNI coordinates (mm)** | | |  |  |
| --- | --- | --- | --- | --- | --- | --- |
| **Cluster size k (mm^3^)** | **Region** | **x** | **y** | **z** | **t-score** | **p-value** |
| **PPMI cohort (n)** |  |  |  |  |  |  |
| 289 | Putamen L | -26 | -4 | -4 | 2.40 | 0.009 |
| **DNA cohort (n)** |  |  |  |  |  |  |
| 416 | Caudate R | 18 | 28 | 4 | 3.58 | <0.001 |
|  | Putamen R | 16 | 18 | -8 | 3.02 | <0.001 |
| 261 | Caudate L | -10 | 24 | -4 | 2.89 | 0.003 |
|  | Putamen L | -14 | 24 | 4 | 2.73 | 0.003 |
| 397 | Pallidum L | -18 | -6 | 0 | 2.79 | 0.004 |
